# Managing the Infodemic: Leveraging Deep Learning to Evaluate the Maturity Level of AI-Based COVID-19 Publications for Knowledge Surveillance and Decision Support

**DOI:** 10.1101/2023.06.20.23291653

**Authors:** Raghav Awasthi, Aditya Nagori, Shreya Mishra, Anya Mathur, Piyush Mathur, Bouchra Nasri

## Abstract

COVID-19 pandemic has taught us many lessons, including the need to manage the exponential growth of knowledge, fast-paced development or modification of existing AI models, limited opportunities to conduct extensive validation studies, the need to understand bias and mitigate it, and lastly, implementation challenges related to AI in healthcare. While the nature of the dynamic pandemic, resource limitations, and evolving pathogens were key to some of the failures of AI to help manage the disease, the infodemic during the pandemic could be a key opportunity that we could manage better. We share our research related to the use of deep learning methods to quantitatively and qualitatively evaluate AI-based COVID-19 publications which provides a unique approach to identify “mature” publications using a validated model and how that can be leveraged further by focused human-in-loop analysis. The study utilized research articles in English that were human-based, extracted from PubMed spanning the years 2020 to 2022. The findings highlight notable patterns in publication maturity over the years, with consistent and significant contributions from China and the United States. The analysis also emphasizes the prevalence of image datasets and variations in employed AI model types. To manage an infodemic during a pandemic, we provide a specific knowledge surveillance method to identify key scientific publications in near real-time. We hope this will enable data-driven and evidence-based decisions that clinicians, data scientists, researchers, policymakers, and public health officials need to make with time sensitivity while keeping humans in the loop.

## 1 INTRODUCTION

The COVID-19 pandemic has highlighted the urgency of strengthening literature-guided disease surveillance to enable policymakers in formulating evidence-driven policies [14][7] [2][17] [9]. The scientific community has rapidly produced many research papers and leveraged artificial intelligence (AI) technologies to understand the virus, its spread, and potential interventions. However, the abundance of AI-based COVID-19 publications necessitates a robust evaluation process to ensure the reliability and usefulness of these works in informing surveillance strategies [4][1],[5] [12][11][15].

This paper presents an innovative approach to enhance disease surveillance capabilities by conducting a qualitative and quantitative evaluation of AI-based COVID-19 publications using deep learning techniques. By harnessing the power of deep learning algorithms, we seek to evaluate the quality, relevance, and impact of research papers in the context of disease surveillance [16][20][13][10].

The outcomes of this study will not only enhance disease surveillance capabilities during the COVID-19 pandemic but also establish a foundation for future surveillance efforts in the face of emerging infectious diseases. By leveraging AI-based deep learning techniques, we can streamline the evaluation process and empower policymakers and public health officials to make informed decisions based on trustworthy and impactful scientific evidence.

Moreover, the implementation of such a maturity model will improve the quality of papers and serve as a catalyst for improving paper quality and industry standards. By following the quality standards established by the maturity model, upcoming research and writing will have great improvements in excellence and merit. Following the global discourse that resulted from the COVID-19 pandemic, more coordination and collaboration can be opened up with the implementation of this new maturity system, which will create a more common understanding and impact of COVID-19 research. In both evaluating current research papers and formulating a structure to follow for upcoming COVID-19 research, this maturity model offers a new step forward in partnership and reliability of research, which is crucial to molding research-based surveillance strategies and interventions.

## 2 METHODS

This paper presents a study that combines a data-driven approach with human evaluation to enhance the robustness, explainability, and real-time relevance of our findings in understanding the maturity patterns of research papers (Fig 1). The level of maturity of a research paper is determined by the answer to the question, ‘Does the output of the proposed model has a significant impact on real-world applications? which is derived from the original article [23], and defined in the context of COVID-19.

**Figure 1.**
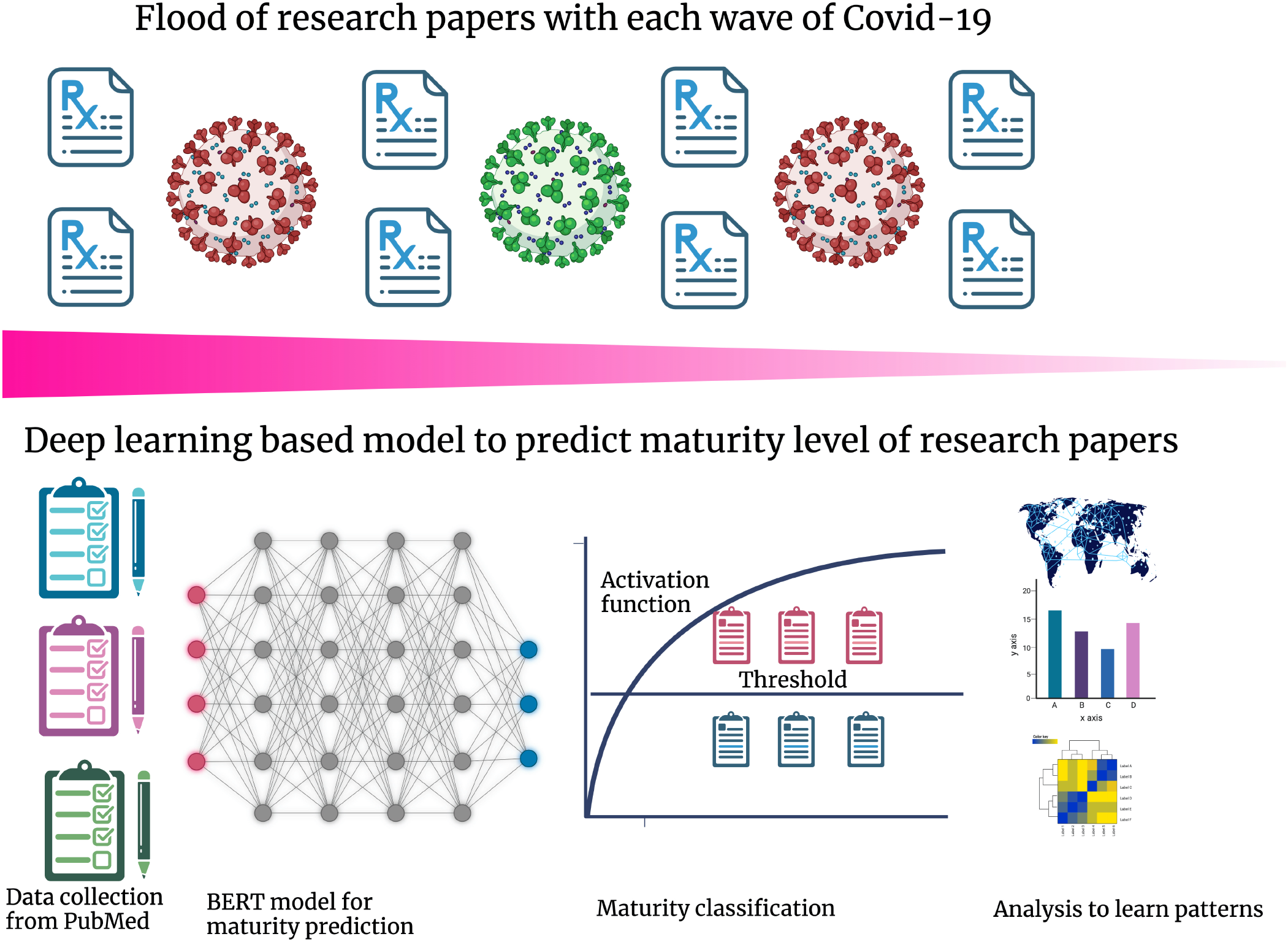
Methodological pipeline for assessing the maturity of COVID-19 research articles. The process involved using peer-reviewed COVID-19 articles from PubMed and determining the country where the senior author based. The BERT neural network model was then employed to assess the maturity level of research articles based on their abstracts. Finally, mature publications were manually annotated with specific details, such as the models and data types utilized in COVID-19 research papers.

### 2.1 Dataset

This study utilized in-house data specifically focused on “Artificial Intelligence in Healthcare” reviews from 2020 to 2022. The data collection process involved conducting a PubMed search using the keywords “machine learning” or “artificial intelligence” in conjunction with the years “2020” “2021” and “2022”. The search was limited to English language publications and those involving human subjects until December 31 of each year. The initial search yielded a preliminary list of 5885, 4164, and 9974 papers for 2020, 2021, and 2022, respectively. Subsequently, each paper underwent an individual examination, and any papers found to have flaws in the PubMed search results or deemed irrelevant to the study were excluded. The final cohort comprised 3232, 2182, and 7916 papers, which were carefully selected, examined, and classified into one or more medical disciplines for the respective years. Among these papers, there were 322, 134, and 504 that specifically focused on Covid-19 [19][3].

### 2.2 Maturity Model

We utilized an approach (Fig. 1) developed to classify the research paper’s maturity based on its abstract [23]. The title and abstract were used as predictors of the paper’s level of maturity. 2, 500 manually labeled abstracts from 1998 to 2020 were utilized to fine-tune the hyperparameters of the BERT PubMed classifier. BERT [8] is a deep learning model for NLP tasks that are built on transformers. BERT’s functioning completely depends on attentional mechanisms that understand the contextual relationships between words in a text. The maturity classifier was validated on a test set (n=784) and prospectively on abstracts from 2021 (n=2494). The test set model had an accuracy of 99% and a precision F1 score of 93%, while the prospective validation model had an accuracy of 99 % and an F1 score of 91%.

### 2.3 Analysis

To calculate the spectrum of maturity in the publication of COVID-19 articles, we have conducted the following analysis:

- First, we calculated the overall year-wise percentage of mature articles.
- Next, we determined the geographical distribution of mature COVID-19 articles regarding the country where the senior author is based.
- Finally, we manually annotated the data type and AI model type used in the matured articles.

## 3 RESULTS

### 3.1 Maturity patterns by the year

In 2020, a total of 322 publications were recorded, out of which 15 were categorized as mature publications. The following year, in 2021, the overall number of publications decreased to 134, with only 2 being classified as mature publications. However, in 2022, the trend reversed, as the total number of publications surged to 504, with 19 being identified as mature publications (Fig. 2A). These results highlight the fluctuating nature of publication output over the three-year period. While there was a significant increase in overall publications from 2021 to 2022, the proportion of mature publications remained relatively low throughout the study period.

**Figure 2.**
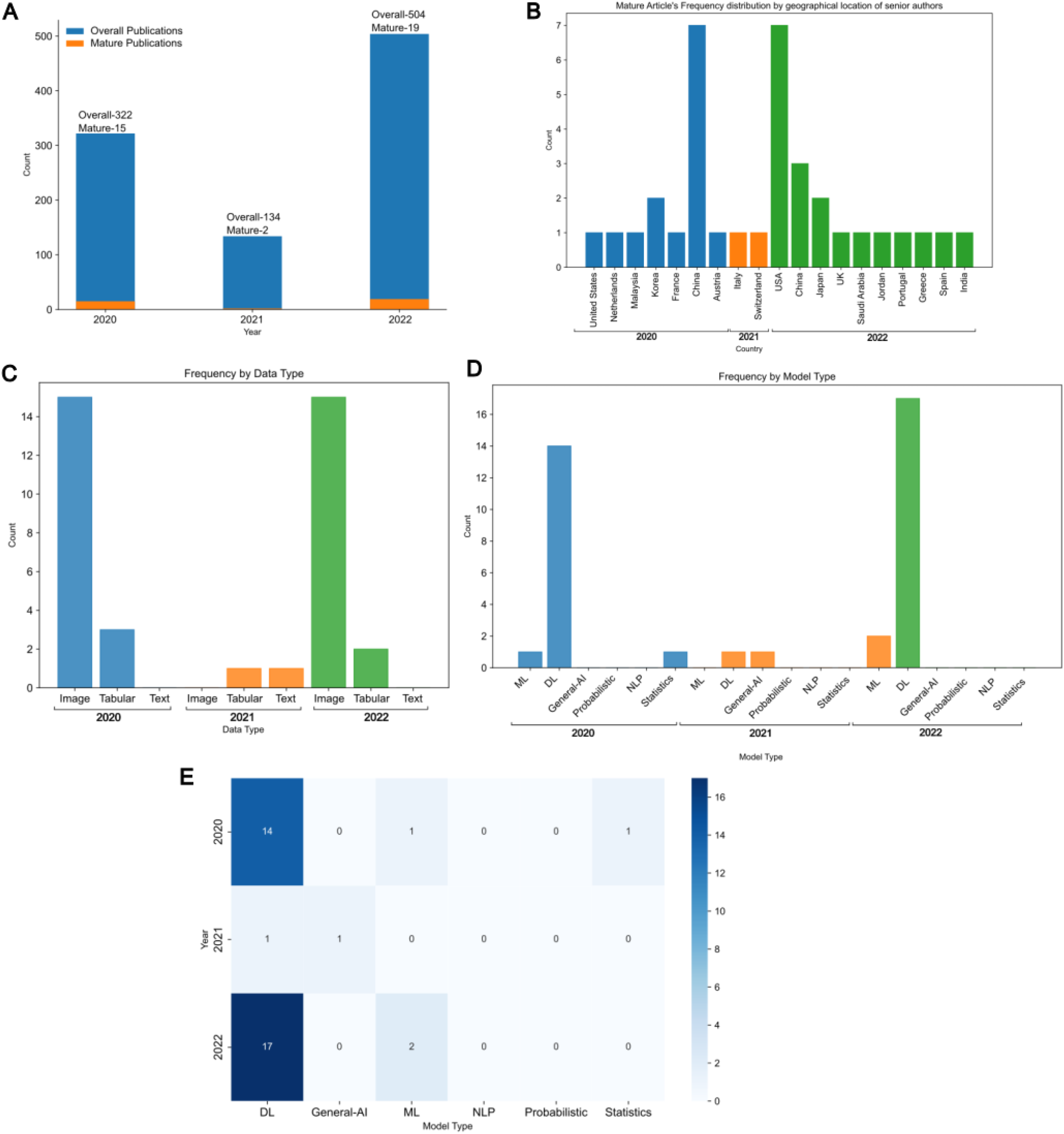
A) Maturity patterns by the year 2020, 2021 and 2022 B) Mature article’s frequency distribution by the geographic location of the senior authors for years 2020, 2021 and 2022 C) Frequency of Mature articles by Data Types (Image, Tabular and Text) for year 2020, 2021 and 2022. D)Frequency of Mature articles by Model Types (ML, DL, General-AI, Probabilistic, NLP, Statistics) for year 2020, 2021 and 2022 E) Heatmap depicting number of Mature articles for all three years classified based on Model type and their comparative analysis.

### 3.2 Mature article’s frequency distribution by the geographic location of the senior authors

Next, we calculated the distribution of mature articles based on the geographic location of the senior authors. The study aimed to analyze the distribution of mature articles across different countries over three years.

In 2020, 15 mature articles were identified, with senior authors from various countries. The United States, the Netherlands, Malaysia, Korea, France, China, and Austria were all represented in the dataset. Notably, China had the highest count with seven mature articles (Fig. 2B).

Moving to 2021, the number of mature articles decreased, and the geographic distribution became more diverse. Italy and Switzerland each had one mature article with senior authors from their respective countries (Fig. 2B). In 2022, the overall count of mature articles increased again. The United States (referred to as the USA in the table) and China continued to dominate, with seven and three mature articles, respectively. Japan, the UK, Saudi Arabia, Jordan, Portugal, Greece, Spain, and India were also represented, each with one mature article (Fig. 2B).

### 3.3 Comparison of Various Datasets and AI Models Employed in Mature Articles

Next, we aimed to analyze the distribution and utilization of different data types and model types in mature articles across various years. The analysis provided valuable insights into the prevalence and trends within the field of mature research. Regarding data types, the findings revealed that image datasets were commonly employed in mature articles, with notable variations across the years. In 2020, 15 articles utilized image datasets, while three articles relied on tabular datasets, and no articles solely focused on textual data. However, in 2021, there was a shift, with no mature articles solely focused on image datasets. Instead, one article utilized a tabular dataset, and another article employed textual data. In 2022, the usage of image datasets increased again, with 15 articles incorporating this type of data, along with two articles based on tabular datasets (Fig. 2C).

Regarding model types, the analysis showed variations in their utilization as well. In 2020, deep learning (DL) models were predominantly employed, with 14 articles utilizing them, while one article used machine learning (ML) models (Fig. 2D, E). There were no mature articles focused on general artificial intelligence (AI), probabilistic models, natural language processing (NLP), or statistical models in that year. In 2021, the distribution shifted, with DL models employed in only one article and one article focusing on general AI. However, no mature articles specifically utilized ML models, probabilistic models, NLP, or statistical models. In 2022, both ML and DL models were employed, with two articles using ML models and seventeen articles utilizing DL models. Nonetheless, there were no mature articles that employed general AI, probabilistic models, NLP, or statistical models in that year (Fig. 2D,E).

## 4 DISCUSSION

In our study, we conducted a qualitative and quantitative evaluation of AI-based COVID-19 publications using deep learning techniques. By harnessing the power of deep learning algorithms, we aimed to evaluate the quality, relevance, and impact of research papers in the context of disease surveillance [18][22][6]. Our objective was to propose a near real-time evaluation of publications that can help guide disease management and further research in the face of emerging infectious diseases with a low level of prior knowledge but rapidly developing new knowledge.

We utilized an in-house dataset compiled from the PubMed data search focusing on the years 2020-2022, with specific keywords related to machine learning and artificial intelligence. The dataset was carefully curated, and papers were classified into medical disciplines to ensure relevance. We then employed a maturity model that utilized the BERT PubMed classifier, a deep learning model for natural language processing (NLP) tasks. The model was fine-tuned using manually labeled abstracts and validated on a test set and prospective abstracts from 2021. Our analysis yielded several important results. First, we observed fluctuations in publication output and the proportion of mature publications over the three-year period. While there was a significant increase in overall publications from 2021 to 2022, the proportion of mature publications remained relatively low. This finding emphasizes the need for rigorous evaluation and selection processes to ensure the reliability and impact of AI-based COVID-19 publications. We also analyzed the mature articles based on geographical location. China and the USA produced aconsistently higher number of mature publications. Other countries exhibit a more varied level of contribution, and the global nature of AI-based research highlights international collaboration. Furthermore, our findings on the data type and model type used in mature articles provided insight into prevailing trends within the field. We found that image data is most commonly used with variation across years. Deep learning models were predominantly employed in the year 2020, but there was a shift in the subsequent years. These findings shed light on the evolving research approaches and the adoption of different data types and model types in the field.

We also examine the fundamental reasons for these discrepancies in publication patterns in addition to analyzing trends in the AI-based COVID-19. First and foremost, data accessibility is important. AI research may benefit from improved access to extensive and varied data-sets in some countries. To process and analyze massive data-sets, this also entails the availability of money, computational infrastructure, and human resources. For instance, the United States has made substantial investments in AI research and infrastructure, including the National Artificial Intelligence Initiative (NAII) and significant funding for cutting-edge research ^1^. Furthermore, each nation has a different capacity for utilizing its resources. It’s possible that certain nations have developed research networks, partnerships, and financing options that support AI development. The regional and temporal heterogeneity seen in our study is influenced by these variables. wWe also recognize the influence of journal specialty areas, special issues, and the accessibility of specialized funding. These elements may encourage or prioritize particular research topics or methodologies, which may have an impact on the publication landscape. The observed variances in AI-based COVID-19 publications may be influenced by such factors. According to various studies, countries such as China, the United States, Japan, the United Kingdom, and Germany are leading the way in AI research and development. However, other countries like Canada, South Korea, France, and Germany are also making significant strides in AI technology. Developing nations like China and India are also rapidly developing their national AI programs [21]. The regional and temporal heterogeneity seen in AI development is influenced by these variables. Additionally, validation and implementation-focused research is hampered by the pandemic’s rapid evolution. The pandemic’s requirement to meet urgent needs may put a time and resource limit on thorough validation and implementation studies. As a result, the published literature can contain a greater percentage of exploratory or preliminary research findings.

However, our work has certain limitations. The exclusion criteria used for selecting papers could introduce bias and limit the generalizability of the findings. The study’s use of deep learning techniques for evaluation may also be subject to limitations, including the potential for model bias and the need for large amounts of labeled data. Finally, It is important to note that these findings are specific to this study and the definition of a mature study. They are also expected in an emergency situation like COVID-19. Our study is also limited to research papers that have been gathered through PubMed and do not include every research paper in AI during the period of study.

In conclusion, our study provides a robust evaluation framework using deep learning techniques for AI-based COVID-19 research publications. The results highlight the disease surveillance capabilities by identifying trends in the publication output, geographical distribution, and research approaches. By providing reliable and impactful scientific evidence, our findings can empower policymakers and public health officials to make informed decisions. We believe our study lays the foundation for future surveillance efforts in the face of emerging infectious diseases, facilitating the development of effective surveillance strategies and interventions.

## Data Availability

All data produced in the present study are available upon reasonable request to the authors

## 5 ACKNOWLEDGMENTS

We acknowledge Graham Dodge, president at PathCheck foundation and Bethany LoMonaco, Director, strategy and development for their constant support. We acknowledge the funding support by Pathcheck foundation awarded to Dr. Aditya Nagori and Mr. Raghav Awasthi. We also acknowledge BrainX for providing the well-annotated datasets.

## Footnotes

1 https://www.ai.gov/strategic-pillars/innovation/

## REFERENCES

[1] AS Albahri, Ali M Duhaim, Mohammed A Fadhel, Alhamzah Alnoor, Noor S Baqer, Laith Alzubaidi, OS Albahri, AH Alamoodi, Jinshuai Bai, Asma Salhi, et al. 2023. A systematic review of trustworthy and explainable artificial intelligence in healthcare: Assessment of quality, bias risk, and data fusion. Information Fusion (2023).

[2] Nisreen A Alwan. 2020. Surveillance is underestimating the burden of the COVID-19 pandemic. The Lancet 396, 10252 (2020), e24.

[3] Raghav Awasthi, Shreya Mishra, Jacek B Cywinski, Kamal Maheshwari, Francis A Papay, and Piyush Mathur. 2023. Quantitative and Qualitative evaluation of the recent Artificial Intelligence in Healthcare publications using Deep-Learning. medRxiv (2023), 2022–12.

[4] Joseph Bullock, Alexandra Luccioni, Katherine Hoffman Pham, Cynthia Sin Nga Lam, and Miguel Luengo-Oroz. 2020. Mapping the landscape of artificial intelligence applications against COVID-19. Journal of Artificial Intelligence Research 69 (2020), 807–845.

[5] Vinay Chamola, Vikas Hassija, Vatsal Gupta, and Mohsen Guizani. 2020. A comprehensive review of the COVID-19 pandemic and the role of IoT, drones, AI, blockchain, and 5G in managing its impact. Ieee access 8 (2020), 90225–90265.

[6] Po-Hsuan Cameron Chen, Craig H Mermel, and Yun Liu. 2021. Evaluation of artificial intelligence on a reference standard based on subjective interpretation. The Lancet Digital Health 3, 11 (2021), e693–e695.

[7] Angel Desai, Pierre Nouvellet, Sangeeta Bhatia, Anne Cori, and Britta Lassmann. 2021. Data journalism and the COVID-19 pandemic: opportunities and challenges. The Lancet Digital Health 3, 10 (2021), e619–e621.

[8] Jacob Devlin, Ming-Wei Chang, Kenton Lee, and Kristina Toutanova. 2018. Bert: Pre-training of deep bidirectional transformers for language understanding. arXiv preprint arXiv:1810.04805 (2018).

[9] Suneela Garg, Nidhi Bhatnagar, Navya Gangadharan, et al. 2020. A case for participatory disease surveillance of the COVID-19 pandemic in India. JMIR Public Health and Surveillance 6, 2 (2020), e18795.

[10] Yu Gu, Robert Tinn, Hao Cheng, Michael Lucas, Naoto Usuyama, Xiaodong Liu, Tristan Naumann, Jianfeng Gao, and Hoifung Poon. 2021. Domain-specific language model pretraining for biomedical natural language processing. ACM Transactions on Computing for Healthcare (HEALTH) 3, 1 (2021), 1–23.

[11] Arash Heidari, Nima Jafari Navimipour, Mehmet Unal, and Shiva Toumaj. 2022. Machine learning applications for COVID-19 outbreak management. Neural Computing and Applications 34, 18 (2022), 15313–15348.

[12] Arash Heidari, Nima Jafari Navimipour, Mehmet Unal, and Shiva Toumaj. 2022. The COVID-19 epidemic analysis and diagnosis using deep learning: A systematic literature review and future directions. Computers in biology and medicine 141 (2022), 105141.

[13] Andreas Holzinger, Peter Kieseberg Edgar Weippl, and A Min Tjoa. 2018. Current advances, trends and challenges of machine learning and knowledge extraction: from machine learning to explainable AI. In Machine Learning and Knowledge Extraction: Second IFIP TC 5, TC 8/WG 8.4, 8.9, TC 12/WG 12.9 International Cross-Domain Conference, CD-MAKE 2018, Hamburg, Germany, August 27–30, 2018, Proceedings 2. Springer, 1–8.

[14] Nahla Khamis Ibrahim. 2020. Epidemiologic surveillance for controlling Covid-19 pandemic: types, challenges and implications. Journal of infection and public health 13, 11 (2020), 1630–1638.

[15] Firuz Kamalov, Aswani Kumar Cherukuri, Hana Sulieman, Fadi Thabtah, and Akbar Hossain. 2023. Machine learning applications for COVID-19: a state-of-the-art review. Data Science for Genomics (2023), 277–289.

[16] Mayara Khadhraoui, Hatem Bellaaj, Mehdi Ben Ammar, Habib Hamam, and Mohamed Jmaiel. 2022. Survey of BERT-base models for scientific text classification: COVID-19 case study. Applied Sciences 12, 6 (2022), 2891.

[17] Patty Kostkova, Francesc Saigí-Rubió, Hans Eguia, Damian Borbolla, Marieke Verschuuren, Clayton Hamilton, Natasha Azzopardi-Muscat, and David Novillo-Ortiz. 2021. Data and digital solutions to support surveillance strategies in the context of the COVID-19 pandemic. Frontiers in Digital Health 3 (2021),707902.

[18] David Lyell, Enrico Coiera, Jessica Chen, Parina Shah, and Farah Magrabi. 2021. How machine learning is embedded to support clinician decision making: an analysis of FDA-approved medical devices. BMJ Health & Care Informatics 28, 1 (2021).

[19] Piyush Mathur, Raghav Awasthi, Kamal Maheshwari, Jacek Cywinski, Vivek Sabharwal, Francis Papay, Ashish Khanna, and Shreya Mishra. 2023. 302: QUANTITATIVE AND QUALITATIVE EVALUATION OF ARTIFICIAL INTELLIGENCE-BASED CRITICAL CARE PUBLICATIONS. Critical Care Medicine 51, 1 (2023), 137.

[20] Marcin Michał Mirończuk and Jarosław Protasiewicz. 2018. A recent overview of the state-of-the-art elements of text classification. Expert Systems with Applications 106 (2018), 36–54.

[21] Neil Savage. 2020. The race to the top among the world’s leaders in artificial intelligence. Nature 588, 7837 (2020), S102–S102.

[22] Jack Wilkinson, Kellyn F Arnold, Eleanor J Murray, Maarten van Smeden, Kareem Carr, Rachel Sippy, Marc de Kamps, Andrew Beam, Stefan Konigorski, Christoph Lippert, et al. 2020. Time to reality check the promises of machine learning-powered precision medicine. The Lancet Digital Health 2, 12 (2020), e677–e680.

[23] Joe Zhang, Stephen Whebell, Jack Gallifant, Sanjay Budhdeo, Heather Mattie, Piyawat Lertvittayakumjorn, Maria del Pilar Arias Lopez, Beatrice J Tiangco, Judy W Gichoya, Hutan Ashrafian, et al. 2022. An interactive dashboard to track themes, development maturity, and global equity in clinical artificial intelligence research. The Lancet Digital Health 4, 4 (2022), e212–e213.

